# Economic and social determinants of health disparities in India: A systematic review of sleep literature

**DOI:** 10.1101/2023.03.13.23287175

**Authors:** Sofia Zoukal, Gabriel Zarate Cordova, Faustin Armel Etindele Sosso

**Affiliations:** Faculté de Médecine et de Pharmacie, Université Hassan II, Casablanca, Maroc; Faculté de médecine de, l’Université Paris-Saclay, Paris, France; Department of Global Health and Ecoepidemiology, Redavi Institute, Montréal, Quebec, Canada

**Keywords:** India, determinant, socioeconomic status, sleep, health disparities, systematic review, public health

## Abstract

Among multiple determinants affecting sleep health, there is people socioeconomic status (SES), a multidimensional concept of an individual’s social, economic and ecological position associated to public health inequalities at different levels. No systematic review on the relation between SES and sleep health has been previously conducted in India. Following Prisma protocol, seven articles were selected. Findings revealed that all studies were cross-sectional. The combined number of participants is N=12,746 participants, composed of 81.15% of adults (n=10,343), 10.56% of children (n=1346) and 8.29% of adolescents (n=1057). The smallest sample was N=268 and the larger was N=7017. The socioeconomic determinants the most reported by authors were perceived SES/composite indices, education, income and employment/occupation. The most reported sleep disturbances were obstructive sleep apnea (OSA), insomnia, restless legs syndrome (RLS) and sleep quality. Higher SES (specifically high education and high income) was associated on one hand in adults, with insomnia and a lower risk for OSA; and on the other hand, in adolescents, with poor quality of sleep and shorter sleep duration. Unemployment was significantly associated with insomnia and risk for pediatric OSA (specifically maternal employment). These findings are coherent with the conceptual socioeconomic model of sleep health published by Etindele Sosso et al. and one previous ecological model of sleep published by Grandner et al., both explaining the relationship between SES and sleep disparities. More studies on the subject and more longitudinal research are necessary to support public health programs related to sleep health disparities in India.

## 1- INTRODUCTION

Health disparities are associated to socioeconomic gradient that can be measured through indicators like education, income, marital status or type of employment [1-7]. These indicators were previously employed in social epidemiology and biomedical research such as those related to cardiovascular system [8], breathing system [9] or sleep mechanisms [1, 2]. They helped established how environments can affect the pathway of an individual’s health status and it was documented extensively that, this relationship was behind a lot of public health issues [4, 6, 10]. Among others important public health issues potentially linked to the social and physical environment, there is sleep health, which is decreasing considerably worldwide since the last decade [10-14].

Sleep is a multifactorial mechanism very sensitive to external inputs with a complex construction at the corner of physiology, sociology, psychology and public health [4, 6, 15]. The concept of sleep health, which is relatively new, promote a multidimensional sleep research’s approach considering a wide range of clinical parameters such as sleep duration, sleep continuity, sleep efficiency or total sleep time [1, 2, 11]; and also no clinical parameters such as sleep quality or sleep insufficiency [12, 16, 17]. Sleep health inequalities represents a public health outcome similar to public health issues previously reported for cardiovascular, mental health and metabolic diseases [18] and among factors influencing variations of these inequalities; socioeconomic status (SES) is one of the most important but strangely also one of the less documented in developing countries [6, 10]. SES is an invisible multidimensional concept of an individual’s social, economic and ecological position associated to public health inequalities at different levels; generated by subjective norms and social ladder defined or adopted by the individual’s community [4, 6, 10, 18-20]. Thus, sleep health disparity is a complex assessment of a socio-ideological and theoretical construct measured in a variety of ways usually considering several determinants such as employment, income, education, occupation and social position [3, 15, 18, 21]. Trends in terms of sleep health disparities seems to be similar everywhere regardless the country [6, 15].

An extensive screening of empirical literature revealed that India was one of the biggest countries with a lack of literature about sleep health relationship with socioeconomic determinants of health disparities. This screening also revealed that, no systematic review on the relation between SES and sleep health has been previously conducted in India. Its pertinent to understand if public health inequalities in terms of sleep observed elsewhere, are the same in this important country with documented variety of national’s health burdens and economic disparities among their multiple ethnocultural populations [22, 23]. The goals of this systematic review is to 1) document socioeconomic determinants of sleep health inequalities in India and 2) recommend future actions and research directions based on evidence.

## 2- METHODS

### 2.1- Literature search

Relevant citations for this review were identified by searching the databases PubMed/Medline and Google scholar between January 2000 and July 2022. A combination of search terms “socioeconomic”, “socio-economic”, ‘‘social position’’, ‘‘social class’’, ‘‘socioeconomic position’’, “determinant*”, “health disparities”, “sleep”, ‘‘sleep disorders’’, ‘‘sleep disturbances’’, ‘‘sleep complains’’, “sleep outcome”, “sleep health” and “India*” was used. All included articles were identified on the basis of relevance to the association between SES determinants and sleep outcomes following the PRISMA guidelines (Fig. 1).

**Figure 1.**
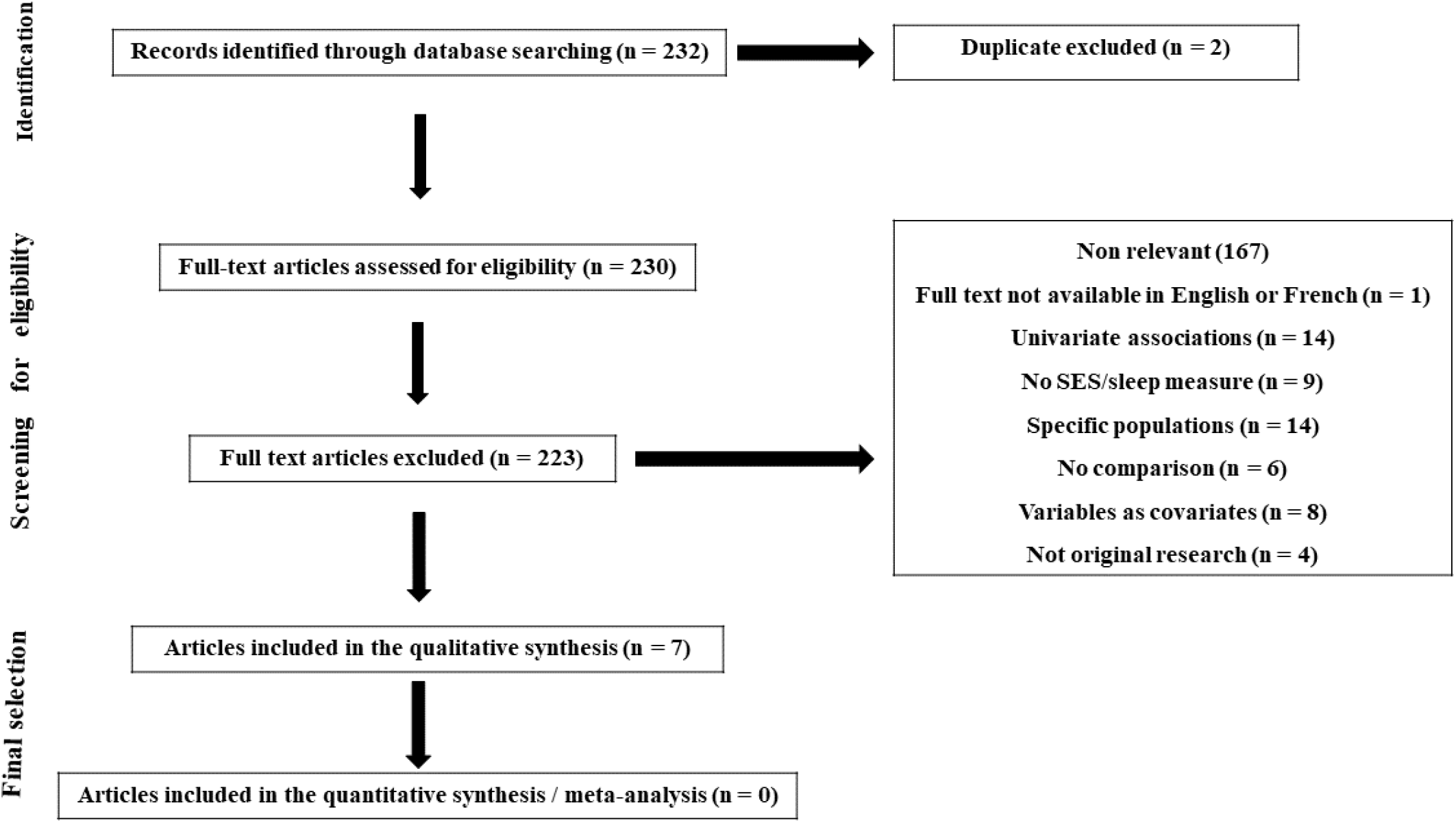
Prisma flowchart of study selection process.

### 2.2- Inclusion and exclusion criteria

Empirical studies were defined as peer-reviewed scientific articles of any design (cross-sectional, retrospective or longitudinal) that assess the relation SES and sleep, including a human sample of any sex, race/ethnicity, gender or age from the general population of India. The study had to include an objective such as education, income, assets, occupation, employment status, perceived SES or a qualitative measure of SES including self-reported items by participants. Aggregate measures of SES (neighbourhood SES or area deprivation indices) were included if participant’s data were not available or reported by authors. For studies with children or adolescents’ participants, perceived family SES measures such as parental education, parental profession or household income were used. Articles were not included excluded when they met one or many of the following criteria: 1) They were reviews or meta-analyses, case series, editorial, case reports, and/or did not present original research, 2) they were not written in English or French, 3) the full text was not available, 4) samples included participants with conditions potentially influence the relation SES and sleep base (for example sleeping pills, chronic sleep disturbances, diseases with sleep symptoms, etc.…), 5) they did not provide statistical significance in cases where the relation between SES indicators and sleep parameters were evaluated.

### 2.3- Quality assessment

The National Institute of Health’s Quality Assessment Tool for Observational Cohort and Cross-Sectional Studies was used to rate the quality of included studies [7]. It assesses 14 quality criteria, asking equal numbers of questions about study objectives, population, exposures, outcomes, follow-up rates, and statistical analysis. Overall quality ratings were calculated by taking the proportion of positive ratings over the sum of applicable criteria. Studies with <50% positive rating were judged as poor quality, 65% as good quality and the rest as fair quality. Complete evaluations criteria of all articles are available in Table 2.

**Table 1.**
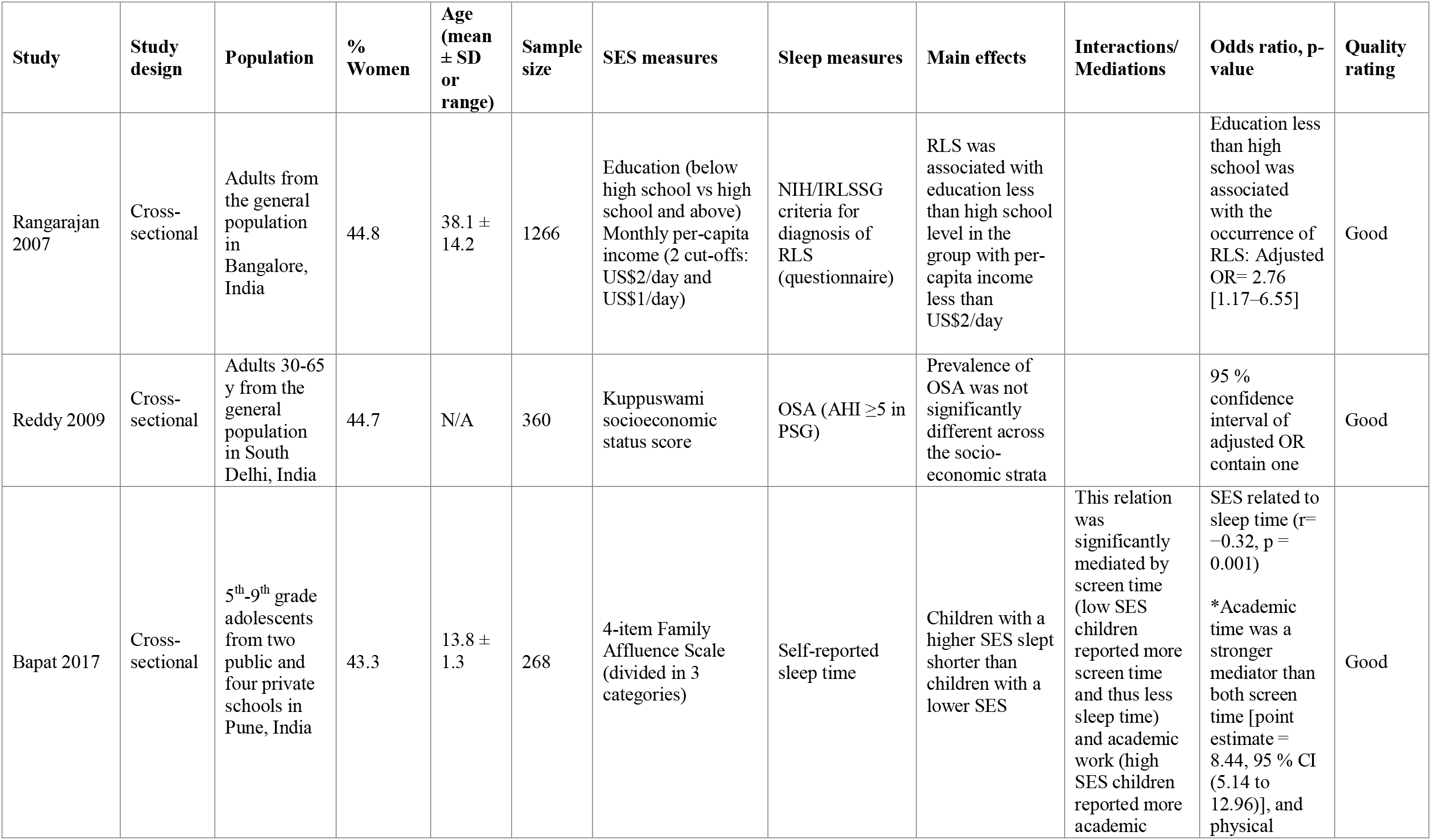
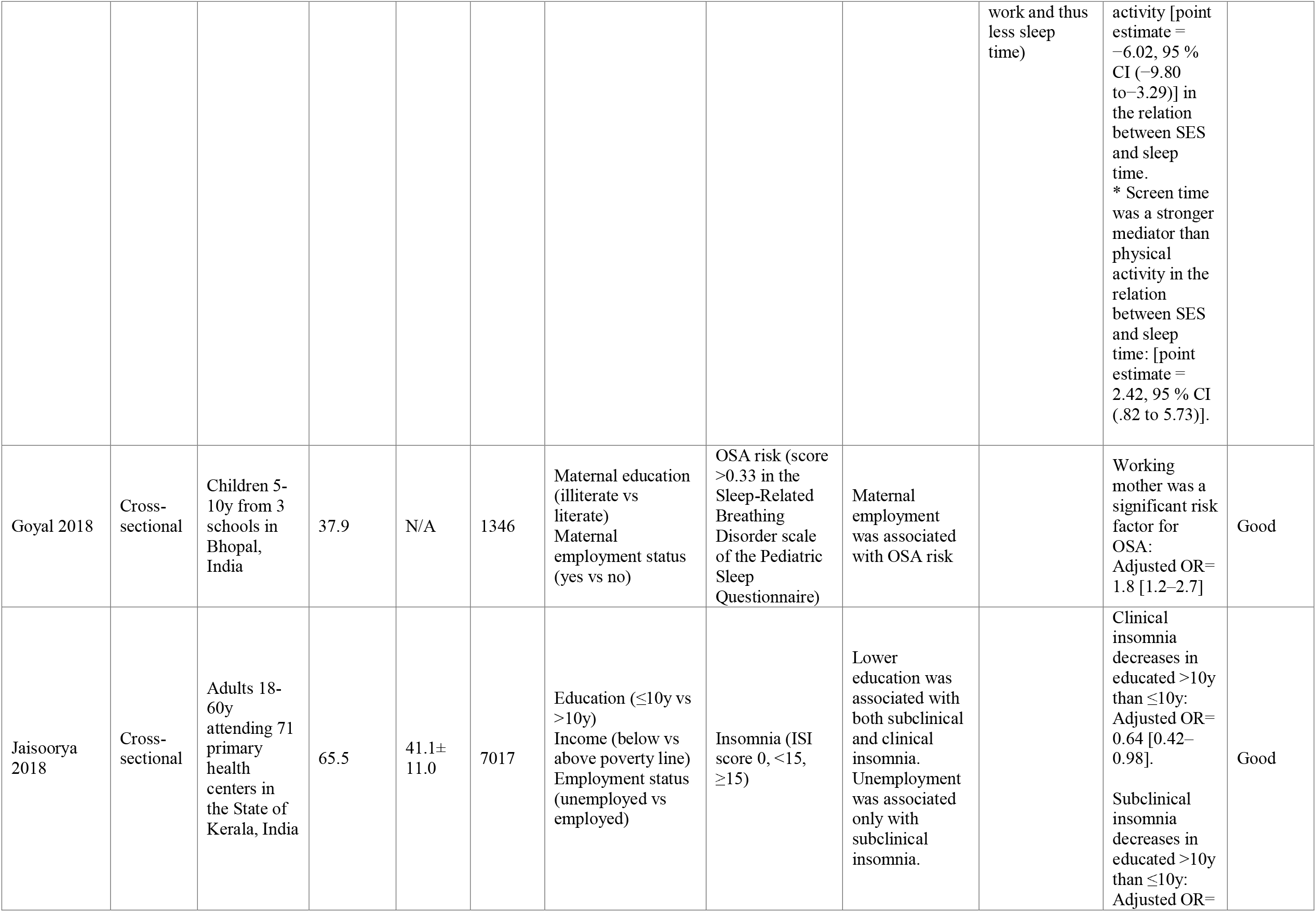
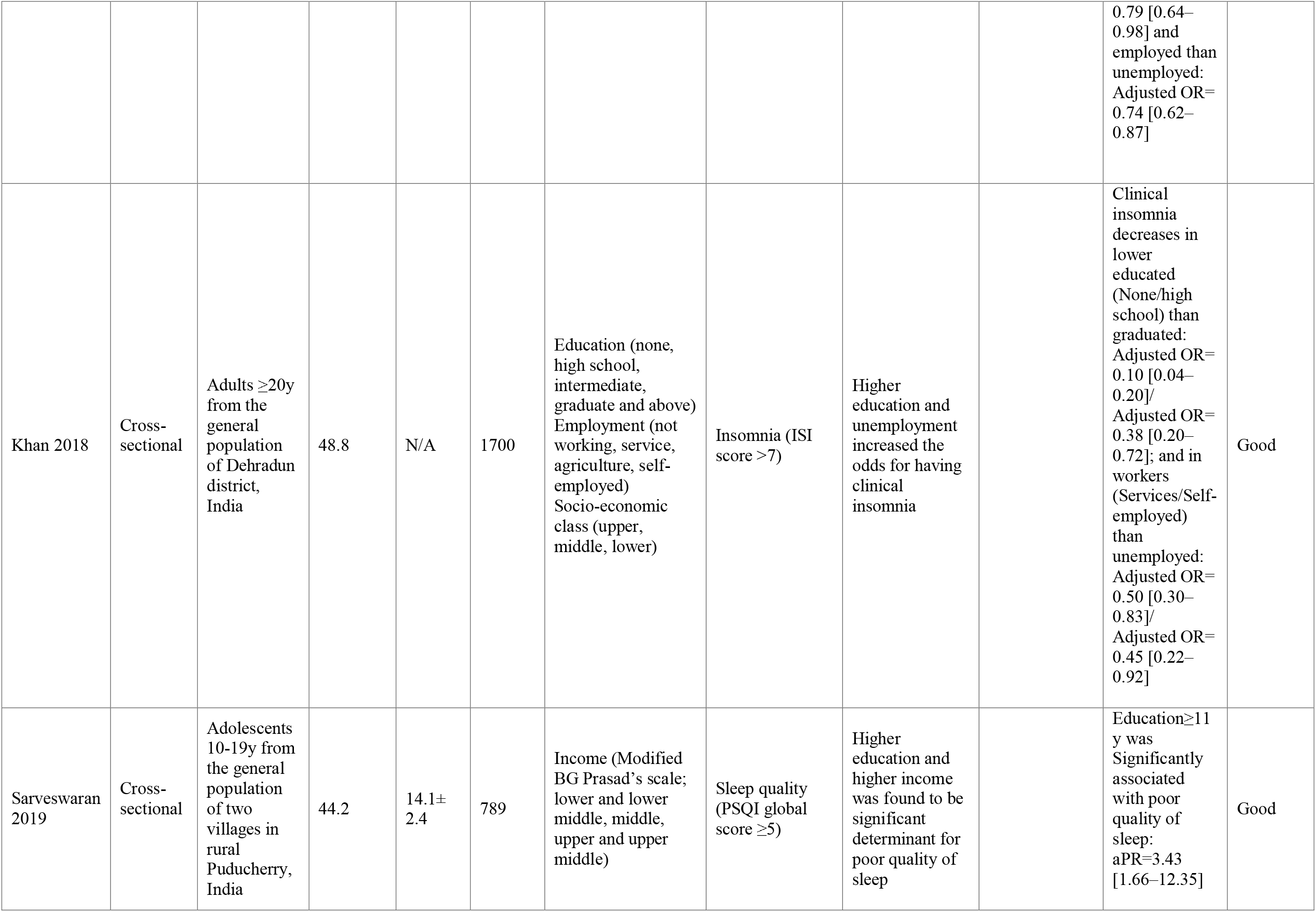
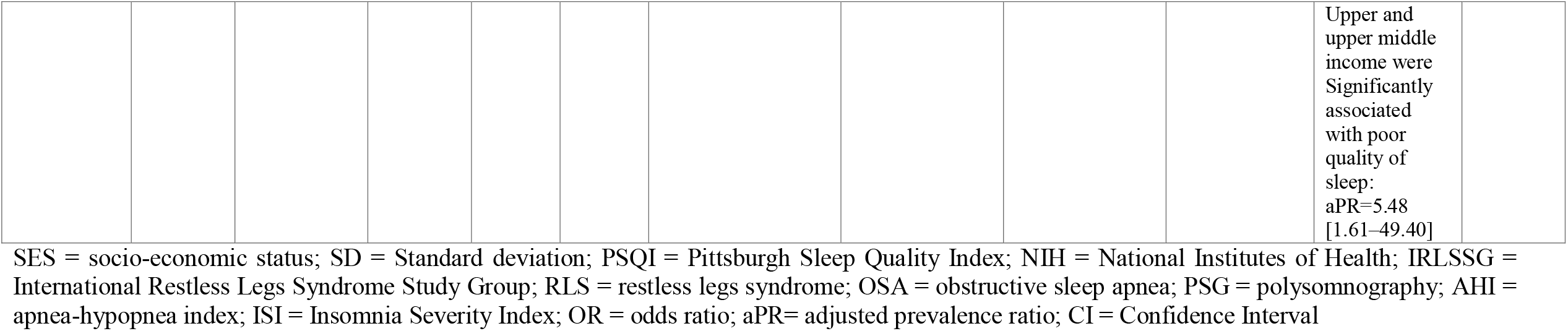
Characteristics of included studies investigating determinants of sleep health disparities in India.

**Table 2.**
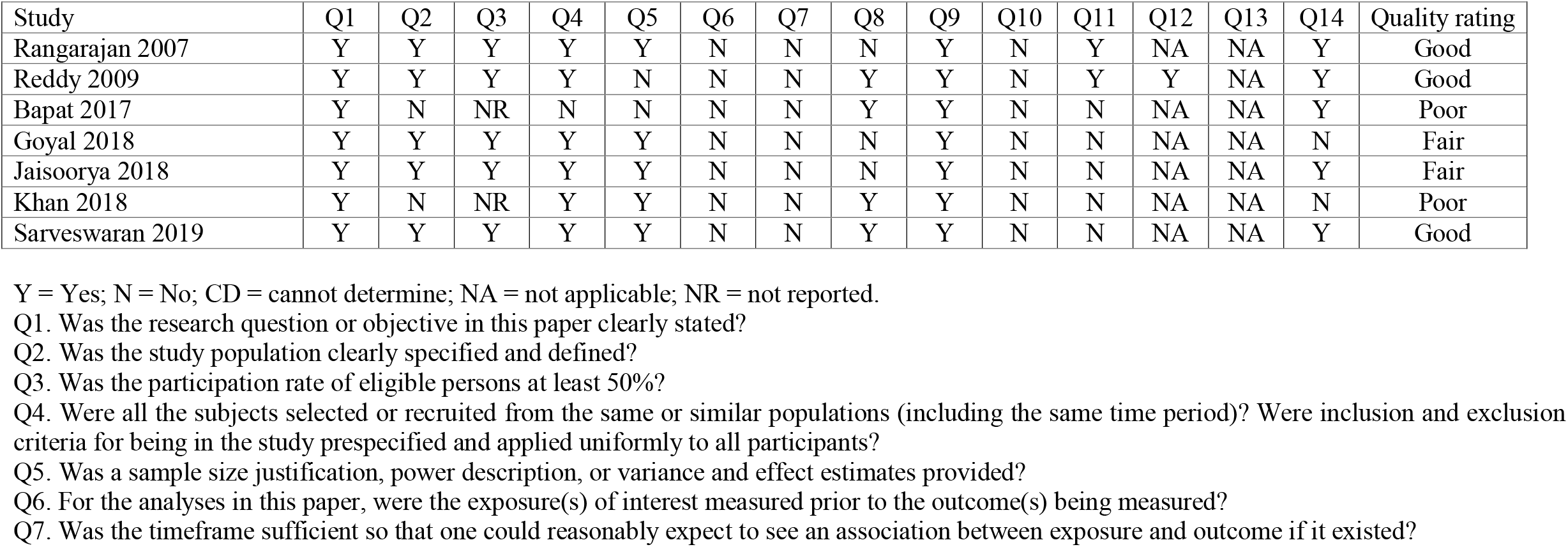
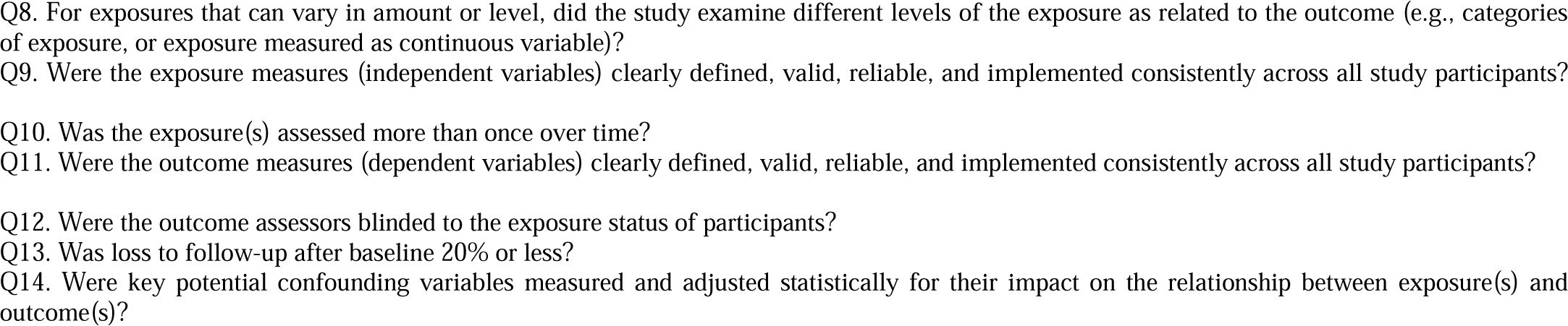
Quality assessment of included studies according to NHLBI’s Quality Assessment Tool for Observational Cohort and Cross-Sectional Studies

## 3- RESULTS

### 3.1- Characteristics of studies selected

Seven articles [24-30] were included in the final sample. All these articles were cross-sectional studies (Table 1) and evaluated as of good quality (Table 2). The combined number of participants is N= 12,746 participants, composed of 81.15% of adults (n = 10,343), 10.56% of children (n = 1346) and 8.29% of adolescents (n= 1057). The smallest sample was N= 268, and the largest was N= 7017. The socioeconomic indicators used were perceived SES/composite indices in three studies [24-26], education in five studies [26-29], income in three studies [27, 29, 30] and employment/occupation in three studies [26, 28, 29].

The measurement instruments and the sleeps disturbances reported were : self-reported sleep time [25], apnea-hypopnea index (AHI) [24] and sleep-related breathing disorder scale (SRBD) [28] for obstructive sleep apnea (OSA), the Insomnia Severity Index (ISI) for insomnia [26, 29], sleep quality using the Pittsburgh Sleep Quality Index (PSQI) [30] and sleep disturbance via National Institutes of Health/International Restless Legs Syndrome Study Group (NIH/IRLSSG) for diagnosis of restless legs syndrome (RLS) [27].

### 3.2- Determinants of sleep health disparities in India

#### Disparities in sleep apnea

In the study by Goyal et al. [28], a significant higher risk for pediatric OSA was observed in association with maternal employment (adjusted odds ratio 1.8; 95% CI: [1.2-2.7]) in school children aged 5-10y. Prevalence of OSA was not significantly different across the updated Kuppuswami socioeconomic status score in the study by Reddy et al. [24] among adults aged 30-65y (*p* > 0.05).

#### Disparities in insomnia

Two studies reported that employment status and education are associated with insomnia (clinical and subclinical) in adults [26, 29]. In the study by Jaisoorya et al. [29], *Subclinical insomnia* was considerably lower in employed (adjusted odds ratio 0.74; 95% CI: [0.62-0.87]) and higher educated >10y (adjusted odds ratio 0.79; 95% CI: [0.64-0.98]). Also, *Clinical insomnia* was considerably higher in unemployed [26, 29]. But for education, prevalence of clinical insomnia decreased with higher educated >10y in the study by Jaisoorya et al. [29] (adjusted odds ratio 0.64; 95% CI: [0.42-0.98]); while it decreased in the lower educated : none (adjusted odds ratio 0.10; 95% CI: [0.04-0.20]) and high school (adjusted odds ratio 0.38; 95% CI: [0.20-0.72]) in the study by Khan et al. [26].

#### Disparities in restless legs syndrome

One study [27] indicated positive association between education and RLS. Occurrence of RLS was significantly associated with education less than high school in the group with higher income cut-off ($2/day) (adjusted odds ratio 2.76; 95% CI: [1.17-6.55]).

#### Disparities in sleep quality

One study [30] showed that education and income was associated with poor quality of sleep among the adolescents. Adolescents with a high educational level > 11*y* (adjusted prevalence ratio 3.43; 95% CI: [1.66 - 12.35]) or a high socio-economic class (adjusted prevalence ratio 5.48; 95% CI: [1.61-49.40]) were more likely to suffer from poor sleep quality.

#### Disparities in sleep duration

Bapat et al. [25] observed a positive association between socioeconomic status and sleep time mediated by academic work mainly and screen time (point estimate = 8.44, 95% CI [5.14 -12.96]), but not by physical activity among adolescents. In the sense that children from a higher SES sleep less as a result of school demands, than children from a lower SES that reporting more screen time which is negatively related to time spent sleeping (*p*=0.001).

## 4- DISCUSSION

### A. Summary of findings

The main findings of the qualitative analyses were as follows: (1) higher SES (specifically education and income) was significantly associated with insomnia and a lower risk of OSA in adults populations, (2) higher SES (specifically education and income) was associated with poor sleep quality and shorter sleep duration in adolescents populations, (3) Unemployment was significantly associated with insomnia, and (4) maternal employment was significantly associated with risk for pediatric OSA.

### B. Relation with current knowledge

The findings of this systematic review are coherent with previous literature including a recent socioeconomic model of sleep health [2, 10, 18] and different socioecological model of sleep [19, 31], stating the strong association existing between the individual socioeconomic status and sleep health. Biological needs for sleep are met by engaging in behaviors that are largely influenced by the environment, social norms and demands, and societal influences and pressures [19]. Understanding the etiology of socioeconomic disparities in sleep could assist public health authorities in preventing the morbidity of socially disadvantaged individuals, in western countries as well in developing countries [32]. Findings of this research supports theories stating that low SES induced sleep disturbances or in other terms, an individual SES is mediating his sleep health following a social gradient measured through markers like education and income [4, 6, 10, 19, 32]. A narrative synthesis of three decades of empirical literature demonstrated that, unhealthy behaviors, increased stress levels and limited access to healthcare in low SES individuals may explain this SES-sleep health gradient [10], because low SES people often reported more sleep disturbances than high SES people. Similarly, it was established that environmental stressors related to climate changes like noise, heat stress and respirable dust are related to an increase of sleep disturbance [33]. In addition, it was showed recently that these disparities are present in several rich countries where social inequities are reduced. For example, a recent systematic review found that in canadian populations, sleep health disparities among children and adolescent are strongly correlated to parental socioeconomic indicators [18]. Findings revealed also that poor parental income, poor family SES and poor parental education are associated with higher sleep disturbances among children and adolescents; same thing with lower education which acts as a predictor of increased sleep disturbances for adults [18]. The same trends were observed with adults and old populations, with low SES associated with high sleep disturbances and low income which was significantly associated with short sleep duration [10, 18]. These results clearly highlight the importance of considering multiple psychosocial and environment risk factors for implementing occupational health and ergonomics interventional programs to prevent sleep disturbances for the entire population, including adolescents and the country’s workforce [33], if governments and employers wish to prevent major expenditures related to inevitable consequences due to an unhealthy sleep [4, 12, 34]. However, the cross-sectional design of most studies related to this relationship and the high heterogeneity in employed measures of SES, reduce the larger promotion of better sleep hygiene and a global standardization of evidence-based policies to improve sleep health of populations across the world [4]. Further research in India is warranted due to important implications for health issues and policy changes.

### C. Recommendations for future research

SES has an unrecognized influence on behavioral risk factors as well as public health strategies related to sleep health disparities. In several countries with a wide range of public health policies and economic challenges, sleep appears to be the main visible consequence of stress induced by difficult living conditions regardless population [13, 35-38]. Obviously, at a more macro level, country’s economic policy influences population’s SES as well as the funding of public health programs. The national and regional public health programs can target directly sleep health, while the same sleep health is affected by stress generated by the individual SES. Thus, SES, economic policy, public health and sleep are linked together. The socioeconomic model of sleep health (Figure 2, Figure 3) developed in previous research [2, 4, 18, 31] may explains all these interconnexions and can be a good start for a more national thinking about the management of Indian’s sleep health. The comparison of sleep health determinants can be made with other diseases determinants (cardiovascular diseases, mental disorders, etc…) to assess the magnitude of their influence, knowing that influence of SES on sleep can be measured objectively and quantitatively [1, 2, 11].

**Figure 2.**
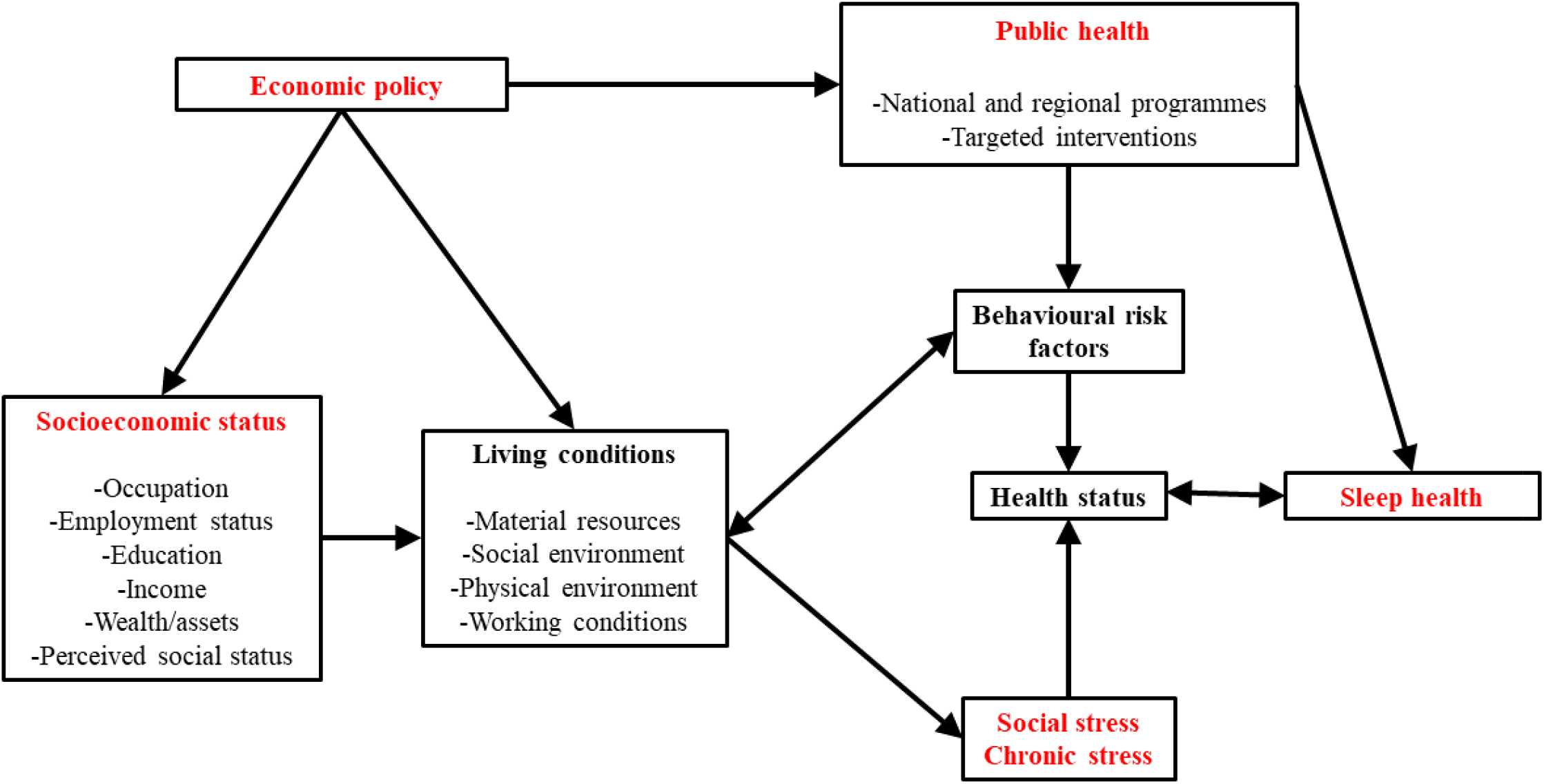
Socioeconomic Model of Sleep Health (adapted with permission from Etindele Sosso FA et al. Eur. J. Investig. Health Psychol. Educ 2022)

**Figure 3.**
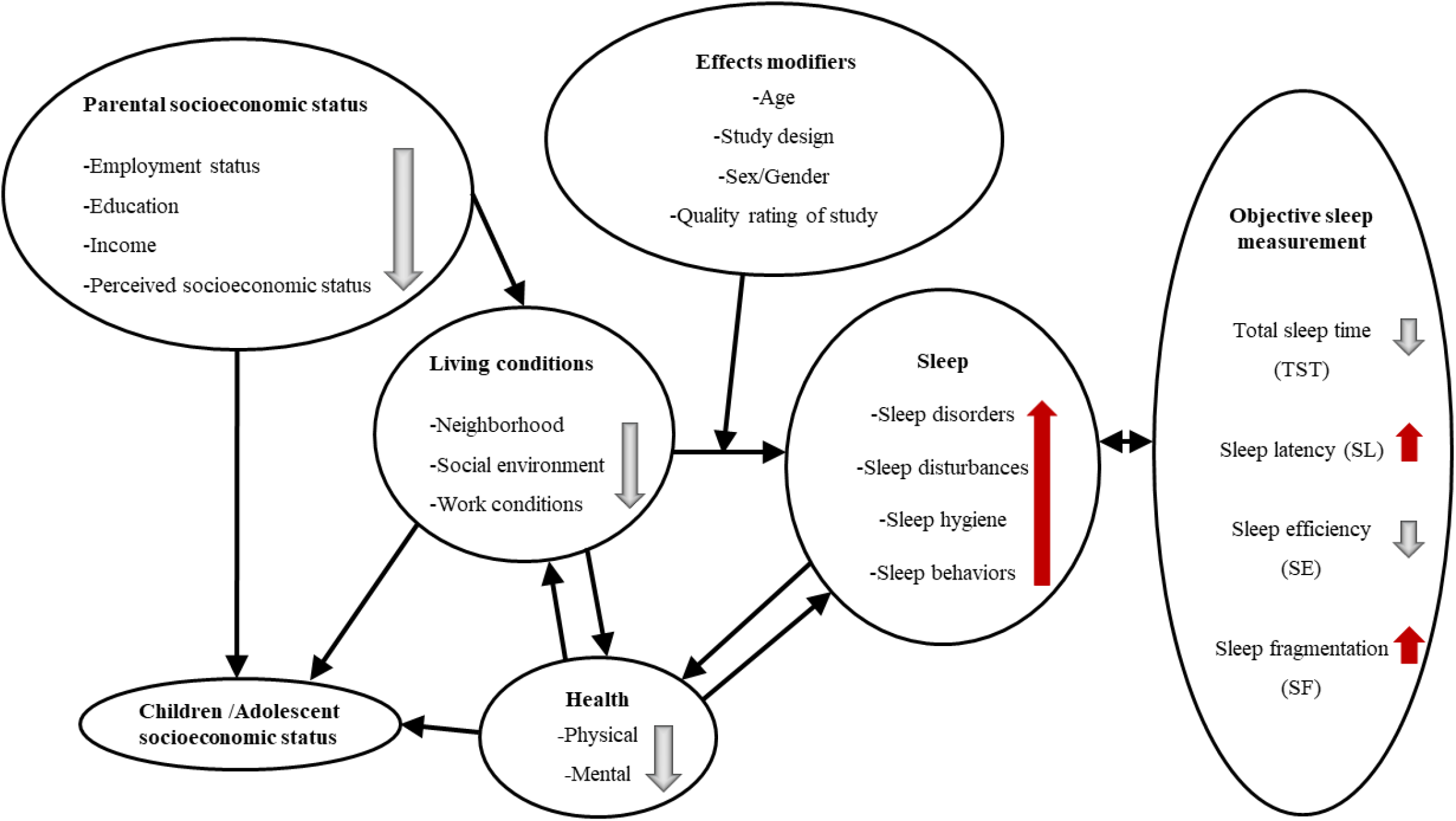
Socioeconomic Model of Sleep (adapted with permission from Etindele Sosso FA et al. Sleep Health 2021)

## Data Availability

All data produced in this present research are already published online and available to everyone.

## Authors’ Statement of Conflict of Interest and Adherence to Ethical Standards

The author reports no conflict of interest.

## References

1. Etindele Sosso, F.A., Measuring Sleep Health Disparities with Polysomnography: A Systematic Review of Preliminary Findings. Clocks Sleep, 2022. 4(1): p. 80–87.

2. Etindele Sosso, F.A., S.D. Holmes, and A.A. Weinstein, Influence of socioeconomic status on objective sleep measurement: A systematic review and meta-analysis of actigraphy studies. Sleep Health, 2021. 7(4): p. 417–428.

3. Papadopoulos, D., et al., Sleep Disturbances Are Mediators Between Socioeconomic Status and Health: a Scoping Review. International Journal of Mental Health and Addiction, 2020.

4. Hale, L., W. Troxel, and D.J. Buysse, Sleep Health: An Opportunity for Public Health to Address Health Equity. Annual Review of Public Health, 2020. 41(1): p. 81–99.

5. Etindele-Sosso, F.A., Insomnia, excessive daytime sleepiness, anxiety, depression and socioeconomic status among customer service employees in Canada. Sleep science (Sao Paulo, Brazil), 2020. 13(1): p. 54–64.

6. Berwick, D.M., The Moral Determinants of Health. JAMA, 2020. 324(3): p. 225–226.

7. Rodriguez, J.M., et al., Social stratification and allostatic load: shapes of health differences in the MIDUS study in the United States. J Biosoc Sci, 2019. 51(5): p. 627–644.

8. Meneton, P., et al., Work environment mediates a large part of social inequalities in the incidence of several common cardiovascular risk factors: Findings from the Gazel cohort. Soc Sci Med, 2018. 216: p.59–66.

9. Petrovic, D., et al., Social inequalities in sleep-disordered breathing: Evidence from the CoLaus|HypnoLaus study. J Sleep Res, 2019. 28(5): p. e12799.

10. Papadopoulos, D. and F.A. Etindele Sosso, Socioeconomic status and sleep health: a narrative synthesis of three decades of empirical research. J Clin Sleep Med, 2022.

11. Wendt, A., et al., Sleep parameters measured by accelerometry: descriptive analyses from the 22-year follow-up of the Pelotas 1993 birth cohort. Sleep Med, 2020. 67: p. 83–90.

12. Holstein, B.E., et al., Difficulties falling asleep among adolescents: Social inequality and time trends 1991-2018. J Sleep Res, 2020. 29(1): p. e12941.

13. Sheehan, C.M., et al., Are U.S. adults reporting less sleep?: Findings from sleep duration trends in the National Health Interview Survey, 2004-2017. Sleep, 2019. 42(2).

14. Mai, Q.D., et al., Employment insecurity and sleep disturbance: evidence from 31 European countries. Journal of sleep research, 2019. 28(1): p. e12763.

15. Grandner, M.A., Chapter 5 -Social-ecological model of sleep health, in Sleep and Health, M.A. Grandner, Editor. 2019, Academic Press. p. 45–53.

16. Wu, W., et al., Sleep quality of Shanghai residents: population-based cross-sectional study. Qual Life Res, 2020. 29(4): p. 1055–1064.

17. Chami, H.A., et al., Sleepless in Beirut: sleep duration and associated subjective sleep insufficiency, daytime fatigue, and sleep debt in an urban environment. Sleep Breath, 2020. 24(1): p. 357–367.

18. Etindele Sosso, F.A., et al., Towards A Socioeconomic Model of Sleep Health among the Canadian Population: A Systematic Review of the Relationship between Age, Income, Employment, Education, Social Class, Socioeconomic Status and Sleep Disparities. European Journal of Investigation in Health, Psychology and Education, 2022. 12(8): p. 1143–1167.

19. Grandner, M.A., Sleep, Health, and Society. Sleep Med Clin, 2020. 15(2): p. 319–340.

20. Phelan, J.C., B.G. Link, and P. Tehranifar, Social conditions as fundamental causes of health inequalities: theory, evidence, and policy implications. J Health Soc Behav, 2010. 51 Suppl: p. S28–40.

21. Ribeiro, A.I., et al., Neighborhood Socioeconomic Deprivation and Allostatic Load: A Scoping Review. Int J Environ Res Public Health, 2018. 15(6).

22. Gupta, I. and S. Mondal, Urban health in India: who is responsible? Int J Health Plann Manage, 2015. 30(3): p. 192–203.

23. Long, X., et al., Exports Widen the Regional Inequality of Health Burdens and Economic Benefits in India. Environ Sci Technol, 2022. 56(19): p. 14099–14108.

24. Reddy, E.V., et al., Prevalence and risk factors of obstructive sleep apnea among middle-aged urban Indians: a community-based study. Sleep Med, 2009. 10(8): p. 913–8.

25. Bapat, R., M. van Geel, and P. Vedder, Socio-Economic Status, Time Spending, and Sleep Duration in Indian Children and Adolescents. J Child Fam Stud, 2017. 26(1): p. 80–87.

26. Khan, I.W., et al., Generalized Anxiety disorder but not depression is associated with insomnia: a population based study. Sleep Sci, 2018. 11(3): p. 166–173.

27. Rangarajan, S., S. Rangarajan, and G.A. D’Souza, Restless legs syndrome in an Indian urban population. Sleep Med, 2007. 9(1): p. 88–93.

28. Goyal, A., et al., Association of pediatric obstructive sleep apnea with poor academic performance: A school-based study from India. Lung India, 2018. 35(2): p. 132–136.

29. Jaisoorya, T.S., et al., Insomnia in primary care-a study from India. Sleep Health, 2018. 4(1): p. 63–67.

30. Sarveswaran, G., et al., Prevalence and determinants of poor quality of sleep among adolescents in rural Puducherry, South India. Int J Adolesc Med Health, 2019. 33(2).

31. Grandner, M.A., Social-ecological model of sleep health, in Sleep and Health, M.A. Grandner, Editor. 2019, Academic Press. p. 45–53.

32. Papadopoulos, D., et al., Sleep Disturbances Are Mediators Between Socioeconomic Status and Health: a Scoping Review. International Journal of Mental Health and Addiction, 2020. 20(1): p. 480–504.

33. Mokarami, H., et al., Multiple environmental and psychosocial work risk factors and sleep disturbances. Int Arch Occup Environ Health, 2020. 93(5): p. 623–633.

34. Wong, K., A.H.S. Chan, and S.C. Ngan, The Effect of Long Working Hours and Overtime on Occupational Health: A Meta-Analysis of Evidence from 1998 to 2018. Int J Environ Res Public Health, 2019. 16(12).

35. Poulain, T., et al., Associations Between Socio-Economic Status and Child Health: Findings of a Large German Cohort Study. Int J Environ Res Public Health, 2019. 16(5).

36. Peltzer, K. and S. Pengpid, Prevalence, social and health correlates of insomnia among persons 15 years and older in Indonesia. Psychol Health Med, 2019. 24(6): p. 757–768.

37. Muller, D., et al., How long do preschoolers in Aotearoa/New Zealand sleep? Associations with ethnicity and socioeconomic position. Sleep Health, 2019. 5(5): p. 452–458.

38. Chami, H.A., et al., Sleepless in Beirut: Sleep Difficulties in an Urban Environment With Chronic Psychosocial Stress. J Clin Sleep Med, 2019. 15(4): p. 603–614.

